# Development of a Novel Risk Prediction Model for Rheumatoid Arthritis–Associated Interstitial Lung Disease (RA-ILD): A Longitudinal Study

**DOI:** 10.64898/2026.06.18.26355441

**Authors:** Yu-ping lv, Si-yi Wang, Hai-ning Piao, Zheng Gong, Ke-qin Zeng, Qiao Zhong, Shu-feng Lei, Min Tong, Wen-yan Ren, Long-fei Wu

## Abstract

**Objectives:** Interstitial lung disease (ILD) is one of the most common and potentially most devastating extra-articular complication of rheumatoid arthritis (RA) and is associated with substantial morbidity and mortality. However, early identification tools remain limited. This study aimed to identify plasma protein biomarkers of RA-ILD and develop an interpretable machine learning model for risk prediction using data from the UK Biobank.

**Methods:** We evaluated the association between baseline RA and incident ILD risk using Cox proportional hazards models, followed by Mendelian randomization (MR) to assess causal relationship. We then analyzed 2,920 plasma proteins (Olink platform) from 781 RA patients. Proteins associated with ILD risk were identified and used to construct eight machine learning models, with performance assessed by ROC and decision curve analysis. The best-performing model was further interpreted using Shapley additive explanations (SHAP) to evaluate feature importance.

**Results:** RA patients had significantly higher ILD risk (HR: 4.425, 95% CI: 3.549-5.518). MR supported a causal association (OR: 1.227, 95% CI: 1.121-1.343). The CatBoost model showed the best performance, achieving an area under the curve (AUC) of 0.884 (95% CI: 0.773,0.996). The SHAP analysis identified LAG3, NPC2, and LAMP3 are the three most important plasma protein predictors of ILD development in patients with RA.

**Conclusion:** Plasma proteomics combined with machine learning may provide a promising approach for identifying biomarkers and predicting ILD risk in patients with RA. LAG3, NPC2, and LAMP3 may serve as candidate biomarkers for RA-ILD and warrant further validation.

**Graphical abstract:** 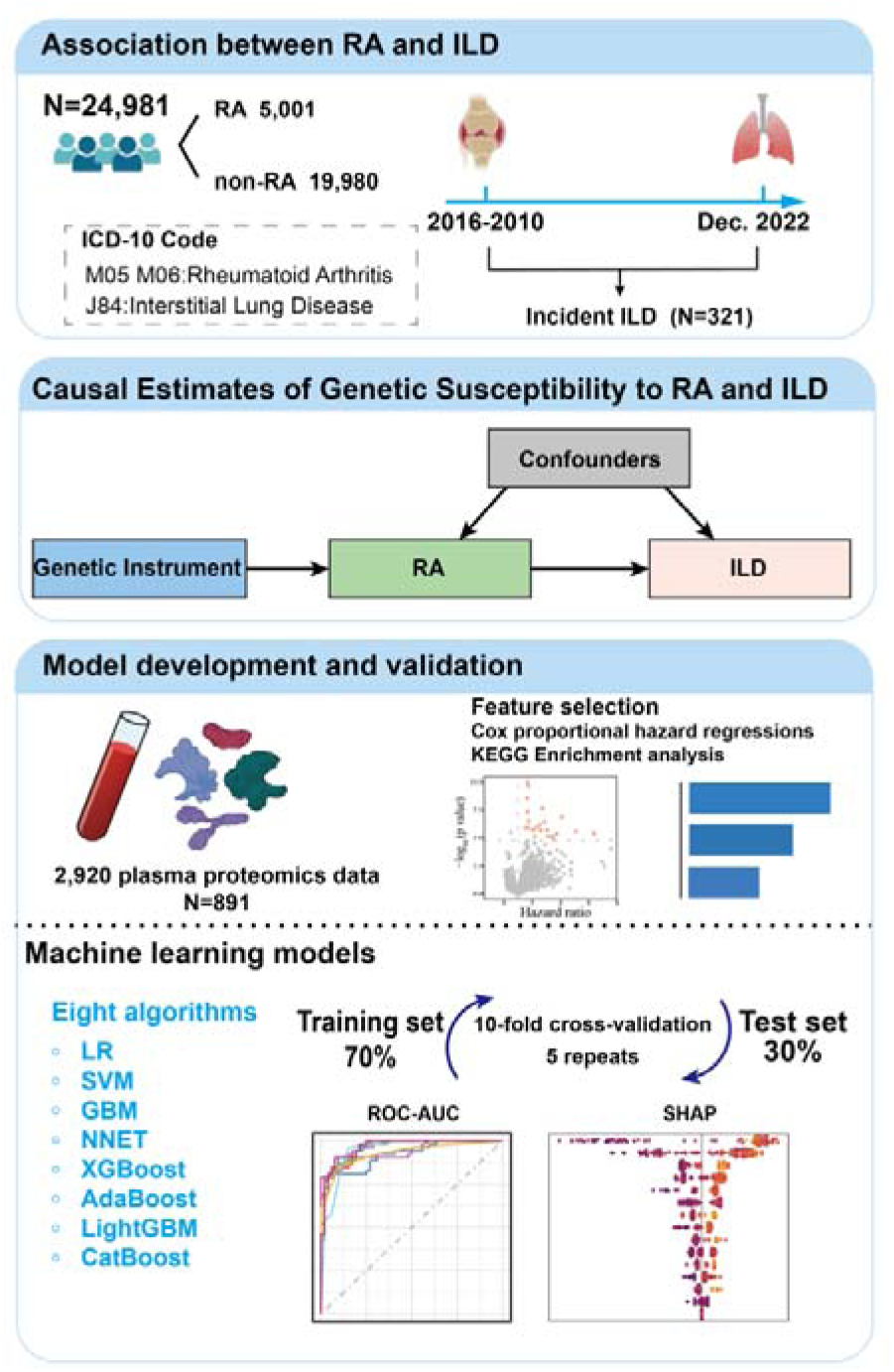

## Introduction

Rheumatoid arthritis (RA) is a common systemic autoimmune disease characterized primarily by chronic tissue inflammation and joint destruction[[1],[2]. Interstitial lung disease (ILD) is a collective term that encompasses a variety of lung conditions characterized by non-infective infiltrates, most commonly in the pulmonary interstitium and alveoli, which in some cases may manifest as structural distortion and irreversible fibrosis[3]. Pulmonary involvement is the most common extra-articular manifestation of RA, and various forms can present as subclinical interstitial lung abnormalities (ILA) or as clinically significant ILD[4]]. RA-associated interstitial lung disease (RA-ILD) significantly contributes to patient morbidity and mortality. The prevalence and incidence of ILD in RA patients depend on the diagnostic methods used and study population selected. Early studies based on chest X-rays estimated the prevalence of ILD to be 5%[5]. A large prospective cohort study of RA patients showed that, over a mean follow-up period of 16.4 years, the estimated prevalence of ILD ranged from 4% to 7.9%, with 10-year, 20-year, and 30-year cumulative incidence rates of 3.5%, 6.3%, and 7.7%, respectively[6]. Clinical risk factors for RA-ILD include older age, male sex, smoking, high RA disease activity, and positivity for anti-citrullinated protein antibodies (ACPA), and rheumatoid factor (RF)[7]-[9].

The early recognition of RA-ILD is a major challenge in clinical practice. In the majority of cases, symptoms of joint inflammation precede the onset of ILD, but in approximately 10% to 20% of cases of RA, interstitial lung changes may be the initial manifestation[10]. Current diagnostic approaches in clinical practice are heterogeneous and primarily symptom-driven. A national survey among rheumatologists revealed that nearly half screen for ILD only in RA patients presenting with respiratory symptoms, utilizing a variable combination of tools such as high-resolution computed tomography (HRCT) (77.6%), X-ray (72.9%), and pulmonary function tests (65.4%)[11]. This lack of standardized screening contributes to diagnostic delays, as conventional imaging often identifies ILD only after significant, irreversible lung damage has occurred[12]. It is therefore crucial for future RA research and treatment to accurately identify the characteristics that predict the development of ILD in RA patients.

To address the limitations of imaging-based diagnosis, research has focused on identifying predictive serum biomarkers. Plasma proteins are involved in immune and cellular processes and can reflect lung injury, fibrosis, and inflammatory pathways. Proteomic analyses have identified candidates like matrix metalloproteinase-3 (MMP-3) and von Willebrand factor (VWF), which, when combined with clinical factors, show promising diagnostic accuracy[13]. Similarly, machine learning models incorporating biomarkers such as Krebs von den Lungen-6 (KL-6), interleukin-6 (IL-6), and cytokeratin 19 fragment (CYFRA21-1) have demonstrated potential for early prediction[12]. However, these studies are typically constrained by cross-sectional designs, relatively small sample sizes, and a limited scope of analyzed proteins, which may restrict the generalizability and robustness of the identified biomarker panels[12], [13]. This study re-evaluates the association between RA and ILD in the general population. Additionally, we utilize Olink plasma protein assay data from participants in the UK Biobank to develop interpretable machine learning models for predicting the future risk of ILD in RA patients, thereby providing a new approach for the early identification of ILD..

## Methods

### Study population

This study utilized data from UK Biobank, a large-scale biomedical database and research resource containing health information on more than 500,000 participants recruited in the United Kingdom between 2006 and 2010[[14]]. At the time of recruitment, participants were aged 40 to 73 years and provided comprehensive baseline data on lifestyle factors, health status, and sociodemographic characteristics[15]. The cohort comprised four assessment phases: (1) baseline assessment conducted from 2006 to 2010, (2) the first follow-up from 2012 to 2013, (3) the second follow-up beginning in 2014, and (4) the third follow-up beginning in 2019. At each stage, participants updated their health information, provided biological samples, and underwent physical measurements. The UK Biobank study received ethical approval from the North West Multicenter Research Ethics Committee (11/NW/0382), and all the participants signed written informed consent forms.

In this study, we established specific inclusion and exclusion criteria for each analysis. For the analysis of the association between RA and ILD, all 502,422 participants from the UKB cohort were included. Exclusion criteria included any of the following: (a) had ILD at baseline, (b) loss to follow-up or withdrawal during the study, (c) death from a non-ILD-related event, or (d) survival time of less than 20 months. The participant selection workflows are shown in **Supplementary Figure 1**. To balance the RA-exposed and control groups via propensity score matching, 24,981 participants were ultimately included. For the association proteomics screening, participants who did not have RA at baseline, those with ILD at baseline, and individuals lacking proteomics data were excluded, resulting in a final sample of 781 subjects.

### RA definitions

RA is defined based on codes M05 or M06 in the International Classification of Diseases, 10th Revision (ICD-10), or code 1464 in self-reported records of non-cancerous diseases.

### Outcome definitions

The main outcomes were the incidence of ILD and survival time; the presence of ILD was defined according to code J84 in ICD-10. Survival time for subjects without ILD was calculated as the time interval between the baseline follow-up date and the last follow-up date (December 31, 2022). For subjects with ILD, survival time was calculated as the time interval between the baseline follow-up date and the date of ILD diagnosis.

### Covariates

The covariates included socio-demographic factors, lifestyle characteristics, and biochemical markers. Sociodemographic covariates included age, sex, race (categorized as White, Mixed, Asian/British Asian, Black/Black British, Chinese/Chinese British, and Others), the Townsend Deprivation Index (TDI), household income, and educational attainment (university degree or higher, non-university degree, and no degree). Lifestyle covariates included body mass index (BMI), smoking status (never smoked, former smoker, or current smoker), alcohol consumption (never drank, former drinker, or current drinker), healthy eating habits, and physical activity. Physical activity was defined as ≥150 minutes of moderate-intensity activity or ≥75 minutes of vigorous-intensity activity per week[16]. The definition and variables used for healthy diet are provided **in the Supplementary Table 1**[17]. Biochemical markers included glycated hemoglobin (HbA1c), high-density lipoprotein (HDL), glucose, and triglycerides.

### Mendelian randomization

To identify causal associations, we conducted a two-sample Mendelian randomization analysis. The genome-wide association study (GWAS) data for RA and ILD were obtained from the IEU Open GWAS Database (https://gwas.mrcieu.ac.uk/). To ensure the reliability and validity of the study results, we employed multiple robust statistical methods, including inverse variance weighting (IVW) under a random-effects model, the weighted median method, Mr. Egger regression, the simple sum of squares method, and the weighted sum of squares method, with the IVW method serving as the primary analysis method[18]. To identify causal associations, we conducted a two-sample Mendelian randomization (MR) analysis. The genome-wide association study (GWAS) data for RA and ILD were obtained from the IEU Open GWAS Database (https://gwas.mrcieu.ac.uk/). To ensure the reliability and validity of the study results, we employed multiple robust statistical methods, including inverse variance weighting (IVW) under a random-effects model, the weighted median approach, the MR-Egger regression, the simple mode, and weighted mode, with the IVW method serving as the primary analysis method [18]. In addition, we assessed the reliability of the study results through sensitivity analyses. We used the Mr-Egger intercept to determine directional horizontal heterogeneity[19]. Subsequently, we applied the Mendelian randomization pleiotropy residual sum and outlier (Mr-PRESSO) test to detect potential horizontal heterogeneity[20]. The Cochrane Q test was used to assess heterogeneity among single-nucleotide polymorphisms (SNPs)[21]. Furthermore, leave-one-out analysis was employed to investigate whether the genetic causal relationship between the exposure and the outcome was influenced by a single SNP. All statistical analyses were performed using the “TwoSampleMR”and“MRPRESSO”packages in R software (version 4.3.2).

### Olink proteomics

The UK Biobank Pharma Proteomics Project (UKB-PPP) conducted a comprehensive study of the plasma proteome. The project utilized the the OlinkTM Explore 3072 Proximity Extension Assay to collect plasma samples from approximately 54,000 UK Biobank participants and perform proteomic analysis. The Olink Explore 3072 comprised eight assay panels covering cardiometabolic, inflammatory, neurological, oncological, cardiometabolic II, inflammatory II, neurological II, and oncological II profiles, detecting a total of 2,923 proteins[22]. The assay methodology used in the UK Biobank study ensured the absence of plate effects, batch effects, or abnormal protein coefficient of variation. For 100% and 99.5% of proteins, the variability attributable to batch effects and plate effects was less than 10%[23]. This study utilized proteomics data from the UK Biobank baseline assessment, which is available at https://biobank.ndph.ox.ac.uk/showcase/label.cgi?id=1839.

### Enrichment analysis

To gain a comprehensive understanding of the biological functions and signaling pathways underlying the significant proteins identified by the two Cox proportional hazards models, we performed the Kyoto Encyclopedia of Genes and Genomes (KEGG). P values were calculated under two-sided tests and statistical significance was defined as a false discovery rate corrected P < 0.05.

### Predictive model development and evaluation

Machine learning (ML) are well-suited for processing large-scale data and can automatically adjust models to optimize performance through techniques such as parameter tuning and cross-validation, this technology has demonstrated outstanding performance in numerous fields[24]-[26]. This study constructed a predictive model for RA-ILD using significant proteins identified via the Cox proportional hazards model. Subsequently, eight machine learning algorithms: Logistic Regression (LR), Support Vector Machines (SVM), Gradient Boosting Machine (GBM), Neural Networks (NNET), Extreme Gradient Boosting (XGBoost), Adaptive Boosting (AdaBoost), Light Gradient Boosting Machine (LightGBM), and Categorical Boosting (CatBoost) were employed for model training and validation. Model performance was evaluated using multiple metrics, including the area under the receiver operating characteristic curve (AUC-ROC), accuracy, sensitivity, specificity, F1 score, calibration curve, decision curve, clinical impact curve, and confusion matrix. The models were interpreted using Shapley additive explanation (SHAP) analysis to elucidate the importance of each feature and the theoretical basis behind the model’s decisions. Statistical analysis was performed using R software version 4.3.2, and a two-sided p-value < 0.05 was considered statistically significant.

## Results

### Baseline characteristics of participants

This study included 467,035 participants, including 5,003 patients with RA and 462,032 patients without RA. **Table 1** shows baseline characteristics by RA status. Compared with patients without RA, those with RA were more likely to be female, older, have a higher BMI, and have higher proportions of smoking and physical inactivity. In addition, participants with RA tended to have lower household income and socioeconomic status. These findings are generally consistent with the known demographic and clinical characteristics of RA populations. During a mean follow-up period of 166.7 months, 2,831 incident cases of ILD were identified. To reduce the impact of potential confounding factors between the RA and non-RA groups and improve comparability, propensity score matching was performed at a ratio of 1:4, matching each participant with RA to four controls without RA. Variables included in the propensity score model were included age, sex, race, income, education level, TDI, BMI, smoking, alcohol consumption, glucose levels, physical activity, and healthy diet. The matched baseline characteristics are shown in **Supplementary Table 2**.

**Table 1.**
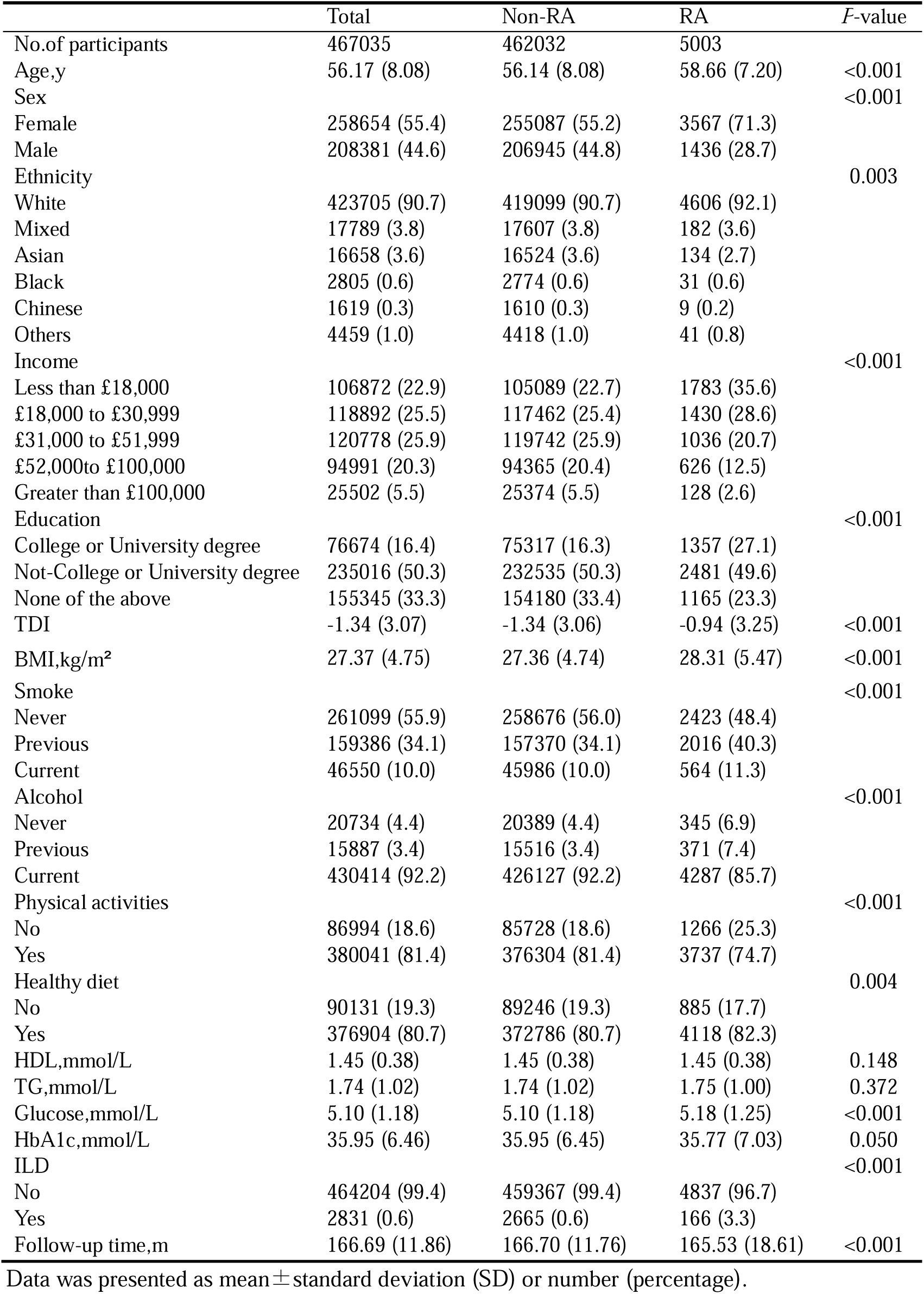
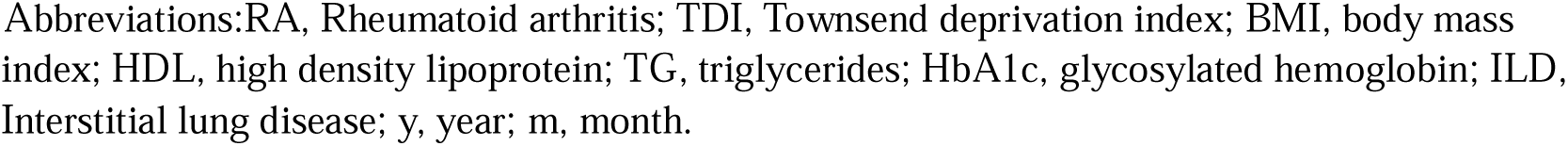
Baseline characteristics of all participants.

|  | Total | Non-RA | RA | P-value |
| --- | --- | --- | --- | --- |
| No.of participants | 467035 | 462032 | 5003 |  |
| Age,y | 56.17 (8.08) | 56.14 (8.08) | 58.66 (7.20) | <0.001 |
| Sex |  |  |  | <0.001 |
| Female | 258654 (55.4) | 255087 (55.2) | 3567 (71.3) |  |
| Male | 208381 (44.6) | 206945 (44.8) | 1436 (28.7) |  |
| Ethnicity |  |  |  | 0.003 |
| White | 423705 (90.7) | 419099 (90.7) | 4606 (92.1) |  |
| Mixed | 17789 (3.8) | 17607 (3.8) | 182 (3.6) |  |
| Asian | 16658 (3.6) | 16524 (3.6) | 134 (2.7) |  |
| Black | 2805 (0.6) | 2774 (0.6) | 31 (0.6) |  |
| Chinese | 1619 (0.3) | 1610 (0.3) | 9 (0.2) |  |
| Others | 4459 (1.0) | 4418 (1.0) | 41 (0.8) |  |
| Income |  |  |  | <0.001 |
| Less than £18,000 | 106872 (22.9) | 105089 (22.7) | 1783 (35.6) |  |
| £18,000 to £30,999 | 118892 (25.5) | 117462 (25.4) | 1430 (28.6) |  |
| £31,000 to £51,999 | 120778 (25.9) | 119742 (25.9) | 1036 (20.7) |  |
| £52,000to £100,000 | 94991 (20.3) | 94365 (20.4) | 626 (12.5) |  |
| Greater than £100,000 | 25502 (5.5) | 25374 (5.5) | 128 (2.6) |  |
| Education |  |  |  | <0.001 |
| College or University degree | 76674 (16.4) | 75317 (16.3) | 1357 (27.1) |  |
| Not-College or University degree | 235016 (50.3) | 232535 (50.3) | 2481 (49.6) |  |
| None of the above | 155345 (33.3) | 154180 (33.4) | 1165 (23.3) |  |
| TDI | -1.34 (3.07) | -1.34 (3.06) | -0.94 (3.25) | <0.001 |
| BMI,kg/m<sup>2</sup> | 27.37 (4.75) | 27.36 (4.74) | 28.31 (5.47) | <0.001 |
| Smoke |  |  |  | <0.001 |
| Never | 261099 (55.9) | 258676 (56.0) | 2423 (48.4) |  |
| Previous | 159386 (34.1) | 157370 (34.1) | 2016 (40.3) |  |
| Current | 46550 (10.0) | 45986 (10.0) | 564 (11.3) |  |
| Alcohol |  |  |  | <0.001 |
| Never | 20734 (4.4) | 20389 (4.4) | 345 (6.9) |  |
| Previous | 15887 (3.4) | 15516 (3.4) | 371 (7.4) |  |
| Current | 430414 (92.2) | 426127 (92.2) | 4287 (85.7) |  |
| Physical activities |  |  |  | <0.001 |
| No | 86994 (18.6) | 85728 (18.6) | 1266 (25.3) |  |
| Yes | 380041 (81.4) | 376304 (81.4) | 3737 (74.7) |  |
| Healthy diet |  |  |  | 0.004 |
| No | 90131 (19.3) | 89246 (19.3) | 885 (17.7) |  |
| Yes | 376904 (80.7) | 372786 (80.7) | 4118 (82.3) |  |
| HDL,mmol/L | 1.45 (0.38) | 1.45 (0.38) | 1.45 (0.38) | 0.148 |
| TG,mmol/L | 1.74 (1.02) | 1.74 (1.02) | 1.75 (1.00) | 0.372 |
| Glucose,mmol/L | 5.10 (1.18) | 5.10 (1.18) | 5.18 (1.25) | <0.001 |
| HbA1c,mmol/L | 35.95 (6.46) | 35.95 (6.45) | 35.77 (7.03) | 0.050 |
| ILD |  |  |  | <0.001 |
| No | 464204 (99.4) | 459367 (99.4) | 4837 (96.7) |  |
| Yes | 2831 (0.6) | 2665 (0.6) | 166 (3.3) |  |
| Follow-up time,m | 166.69 (11.86) | 166.70 (11.76) | 165.53 (18.61) | <0.001 |

Data was presented as mean ± standard deviation (SD) or number (percentage).
Abbreviations:RA, Rheumatoid arthritis; TDI, Townsend deprivation index; BMI, body mass index; HDL, high density lipoprotein; TG, triglycerides; HbA1c, glycosylated hemoglobin; ILD, Interstitial lung disease; y, year; m, month.

### Association between RA and ILD

Cox proportional hazards regression analysis was performed using the propensity score-matched cohort. The association between RA and the incidence of ILD is shown in **Table 2**. In the multivariable-adjusted Cox proportional hazards model, RA was significantly associated with an increased risk of ILD (HR: 4.425, 95% CI: 3.549, 5.518) compared with participants without RA. In addition, male gender, older age, and smoking were all significantly associated with an increased risk of ILD, whereas physical activity was associated with a lower risk of ILD development (**Supplementary Figure 2**).

**Table 2.**
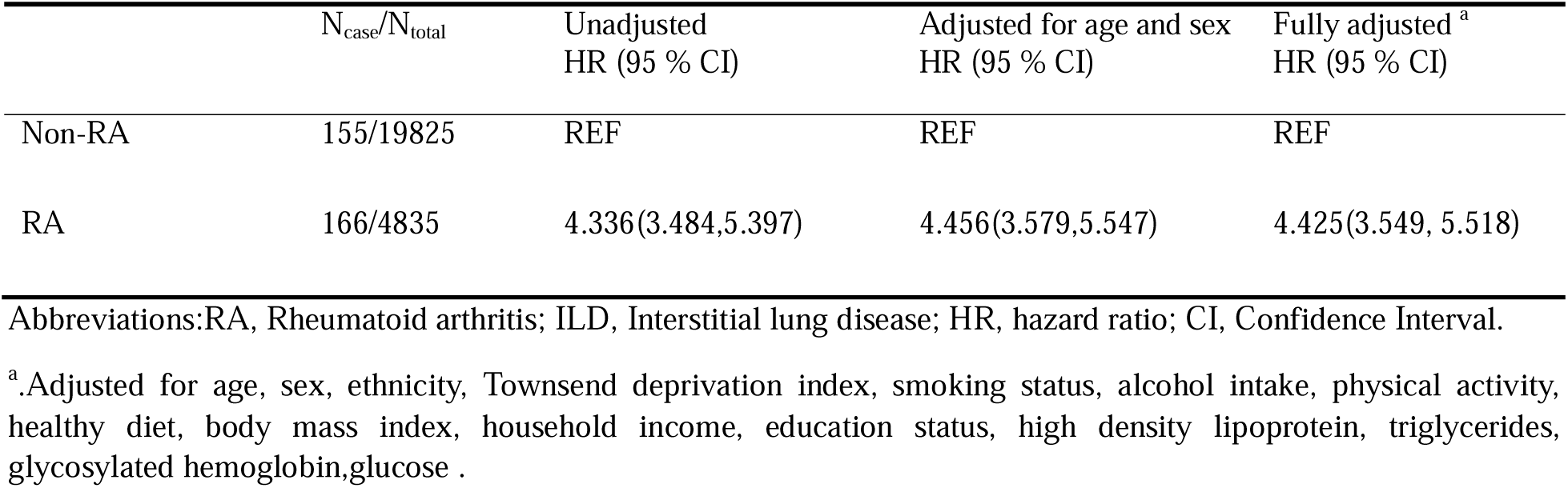
Associations of RA with ILD.

### Causal Estimates of Genetic Susceptibility to RA and ILD

To satisfy the key assumption of MR that the instrumental variables (IV) are strongly correlated with the exposure, we first identified single-nucleotide polymorphisms (SNPs) associated with RA at the genome-wide level (p < 5×10 ). All selected IVs had F-statistics exceeding 10, indicating a low risk of weak instrument bias. Linkage disequilibrium (LD) analysis using a genetic distance threshold of 10,000 kb and an r ² threshold of 0.001 to ensure the independence of the selected SNPs. Palindromic SNPs were excluded. Finally, 24 RA-associated SNPs were retained as IVs for subsequent MR analyses (**Supplementary Table 3**).

Using the IVW method, genetically predicted RA was significantly associated with an increased risk of ILD with an odds ratio (OR) of 1.227 (95% CI: 1.121,1.343, P = 9.83×10^-6^) (**Table 3**). Sensitivity analyses further supported the robustness of this finding. Although Cochran ’ s Q test indicated heterogeneity among the SNP-specific estimates (Q-pval = 0.007), the MR-Egger intercept test showed no evidence of horizontal pleiotropy (p =0.371). Moreover, leave-one-out analysis did not identify any single SNP that substantially influenced the overall estimate (**Supplementary Figure 3**).

**Table 3.** MR results for the causal relationship between RA and ILD.

| Exposure | Outcome | Method | nSNP | $\beta$ | SE | OR(95%CI) | P |
| --- | --- | --- | --- | --- | --- | --- | --- |
| Rheumatoid arthritis | Interstitial lung disease | MR Egger | 24 | 0.155 | 0.072 | 1.167(1.015,1.343) | 0.042 |
|  |  | Weighted median | 24 | 0.152 | 0.046 | 1.164(1.063,1.274) | 0.001 |
|  |  | IVW | 24 | 0.204 | 0.046 | 1.227(1.121,1.343) | <0.001 |
|  |  | Simple mode | 24 | 0.282 | 0.083 | 1.326(1.127,1.560) | 0.002 |
|  |  | Weighted mode | 24 | 0.163 | 0.045 | 1.177(1.078,1.286) | 0.001 |

Abbreviations:RA, Rheumatoid arthritis; ILD, Interstitial lung disease; MR, Mendelian randomization; IVW, inverse variance weighted; nSNP, number of single nucleotide polymorphisms; OR, odds ratio; CI, Confidence Interval.

### Development and Validation of Machine Learning Predictive Models

Among the 2,920 proteomic biomarkers tested, Model 1 identified 23 proteins significantly associated with the onset of ILD after adjusting for age, sex, and race(**Figure 1a**). Subsequently, we performed a sensitivity analysis using Model 2, which additionally adjusted for BMI, smoking, alcohol consumption, and TDI; this model identified 20 proteins significantly associated with the onset of ILD (**Figure 2b**). A total of 19 proteins that were consistently significant in both models were selected for subsequent machine learning model construction. All 19 proteins were positively associated with the risk of ILD. After Bonferroni correction, GADD45B and PLAU showed the strongest associations with ILD risk, with hazard ratios of 1.729 and 1.873, respectively. To characterize the biological properties and regulatory pathways of proteins associated with RA-ILD, we performed a functional enrichment analysis. The results of the enriched biological pathways are shown in the **Figure 1c**, including cytokine-cytokine receptor interactions, interactions between viral proteins and cytokines and cytokine receptors, and the NF-κB signaling pathway.

**Figure 1.**
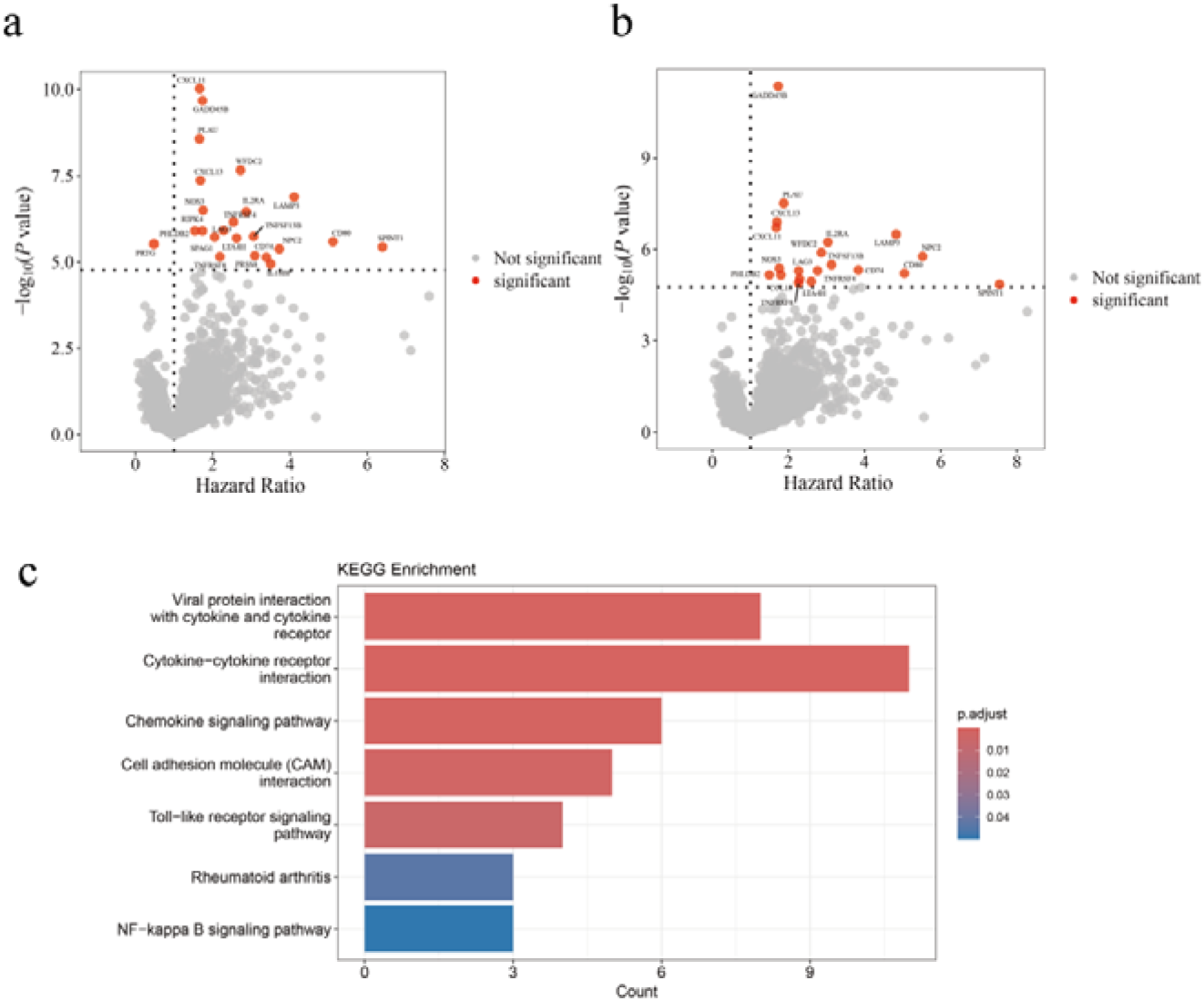
Associations of plasma proteins with ILD. Volcano plots showing the HR (x axis) and −log10(P value) (y axis) for the global associations of 2,920 proteins with ILD. All results for both Cox proportional hazard regression models 1 and 2 are shown here. **a.** Model 1 was adjusted for age, sex, and ethnicity. **b.** Model 2 was additionally adjusted for BMI, smoking, alcohol consumption, and TDI. c.Enrichment for Kyoto Encyclopedia of Genes and Genomes (KEGG) pathways enrichment analysis for the 19 selected proteins.

**Figure 2.**
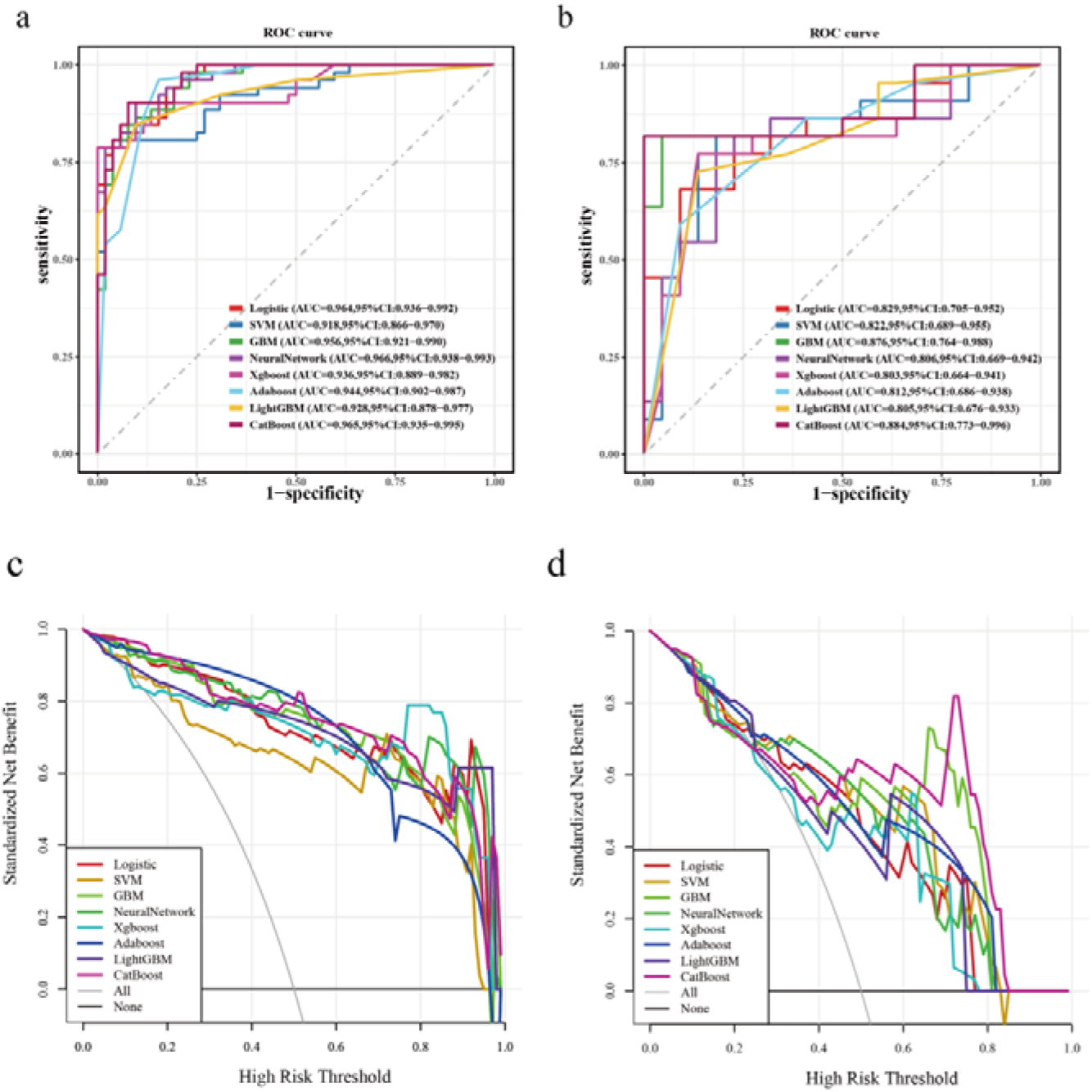
Performance evaluation of the model in the training and test sets. **a.**ROC curve of the training set. **b.**ROC curve of the test set. **c.**DCA curve of the training set and test set. **d.**DCA curve of the training set and test set.

The dataset was randomly split into a training set (70%, n=547) and a validation set (30%, n=234). Because of class imbalance in the original dataset, synthetic minority oversampling technique (SMOTE) was applied to the training set. Eight machine learning models, including LR, GBM, NN, XGBoost, AdaBoost, LightGBM, CatBoost, and SVM, were used to predict the risk of ILD in patients with RA. Model performance was evaluated using accuracy, AUC-ROC, decision curve analysis (DCA), precision and F1 score. Among all the models, CatBoost model demonstrated the best performance (**Figure 2**), with an AUC of 0.884 on the training set and 0.965 on the test set. Therefore, the CatBoost model was selected as the final predictive model.

SHAP analysis was used to interpret the CatBoost model and quantify the contribution of each protein to ILD risk prediction. SHAP bar plots and beeswarm plots showed that LAG3, NPC2, and LAMP3 were among the most influential predictors in the model (**Figure 3a,3b**). Higher LAG3 levels were associated with increased predicted ILD risk, whereas lower LAG3 levels were associated with reduced predicted risk.

**Figure 3.**
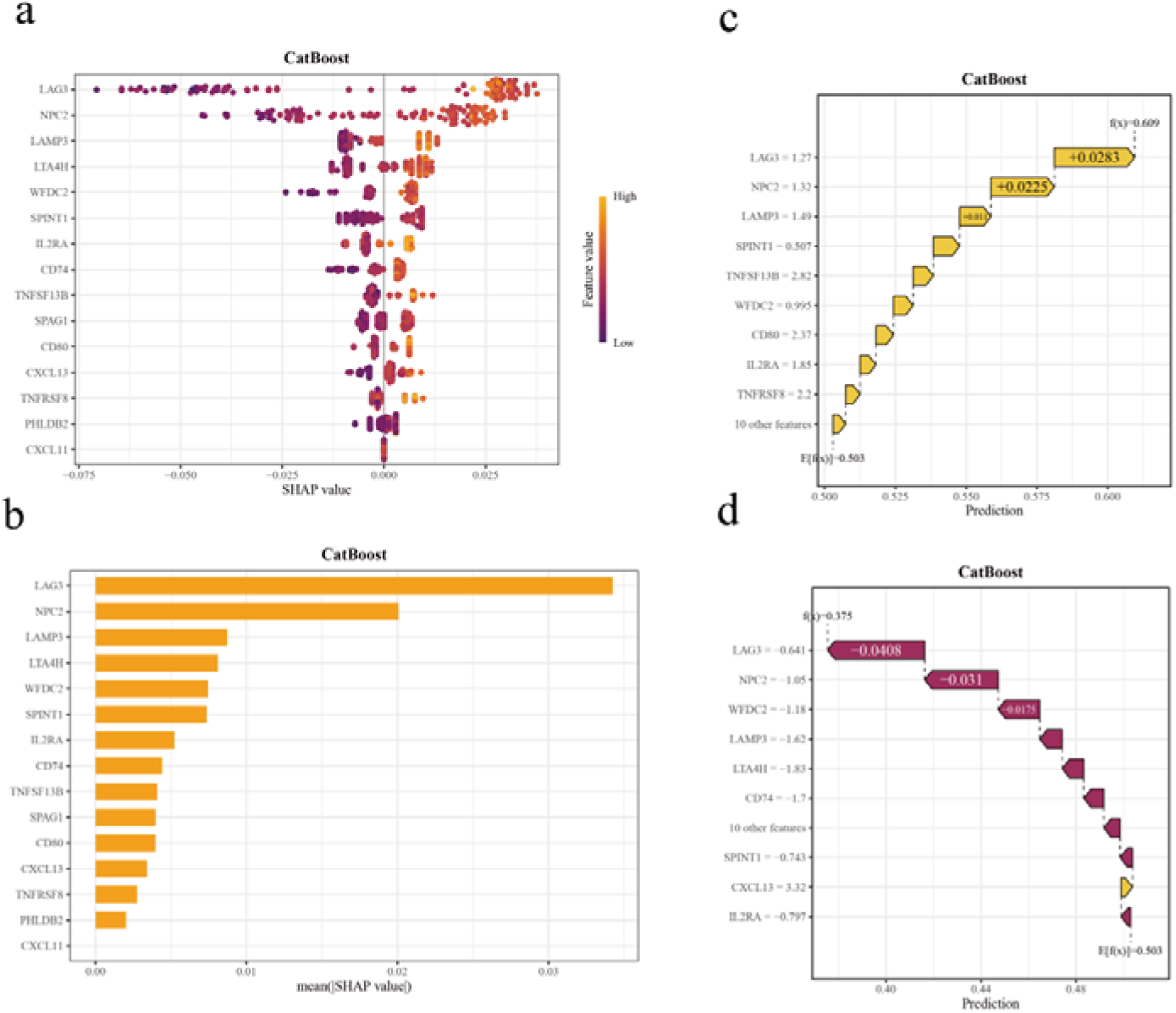
Interpretability analysis of the CatBoost model. **a.**SHAP beeswarm plots of CatBoost model features. **b.**SHAP bar plots of CatBoost model features. **c.**SHAP waterfall plots of the RA-ILD patient. d.SHAP waterfall plots of the RA patient without ILD.

At the individual prediction level, SHAP waterfall plots were generated to explain how increases or decreases in specific feature values lead to an increase or decrease in the risk of the outcome, thereby explaining the source of the model’s prediction for that sample. In the patient with ILD, LAG3, NPC2, and LAMP3 increased the predicted risk, with SHAP values of 0.0283, 0.0225, and 0.0110, respectively. The final predicted risk score was 0.609, exceeding the baseline predicted risk of 0.503 (**Figure 3c**). Conversely, in the patient without ILD, LAG3, NPC2, and WFDC2 contributed to a lower predicted risk, with SHAP values of -0.0408, -0.0310, and -0.0175, respectively. The final predicted risk score was 0.375, which was lower than the baseline predicted risk (**Figure 3d**).

## Discussion

This study first utilized large-scale data from public databases to identify a significant association between RA and ILD; RA was found to be associated with an increased risk of developing ILD, and this association remained consistent after adjusting for covariates. Consequently, evidence of a causal relationship between RA and ILD was provided. Finally, we conducted large-scale proteomics analysis using eight machine learning algorithms to construct a model comprising 19 proteins, enabling the non-invasive prediction of future ILD risk in RA patients. We further employed SHAP analysis to interpret and visualize the CatBoost model, thereby enhancing its clinical utility.

ILD is a serious complication of RA. Previous studies have shown that RA-ILD patients have a significantly higher mortality rate than patients with RA without ILD[27]. The median survival reported in previous studies on RA-ILD ranges from 3 to 10 years. Early identification and intervention are crucial for improving prognosis. Known risk factors for RA-ILD include smoking, male gender, exposure to pollutants, and high levels of anti-citrullinated protein antibodies[28]. Using the CatBoost model, this study screened for several key plasma proteins associated with ILD in RA patients, ranked by Shapley histogram as follows: LAG3, NPC2, LTA4H, SPINT1, LAMP3, WFDC2, IL2RA, TNFRSF8, CD80, CD74, CXCL13, and others.

LTA4H is a key enzyme involved in the inflammatory cascade associated with the arachidonic acid pathway, and neutrophil-mediated inflammatory responses are commonly observed in various pulmonary diseases, such as acute lung injury (ALI) or interstitial pneumonia (IPF)[29]. Experimental studies[30] have shown that the activation of LTA4H ultimately influences the conversion of LTA4 into the pro-inflammatory mediator leukotriene B4 (LTB4) or the anti-inflammatory leukotriene A4 (LXA4). Both excessive or insufficient LTA4H enzymatic activity directly affect susceptibility to severe diseases. LTB4 is one of the most potent chemotactic agents for neutrophils, and its abnormal increase is associated with excessive inflammation; in such cases, the disease is primarily caused by immunopathology. Previous studies have demonstrated [31]-[33] that the use of LTA4H inhibitors in mouse models of ALI and IPF significantly attenuates early neutrophil-mediated inflammatory responses. Although further validation is needed to determine whether LTA4H inhibitors can serve as lead compounds for the treatment of IPF and ALI, they offer a potential therapeutic target for patients with ALI and IPF. Furthermore, LTB4 has been shown to play a significant role in the inflammatory response in COPD patients, with this mediator being upregulated in this population[34]. Results from a population-based study indicate a potential association between the LTA4H rs1978331C variant (in exon 11) and lung function parameters — specifically forced expiratory volume in one second (FEV1) and the FEV1/FVC ratio—in smokers[35].

LAMP belongs to a family of highly glycosylated lysosomal proteins. They account for approximately half of the lysosomal membrane proteins and form a glycoprotein coat on the inner surface of the lysosomal membrane. LAMP3, recently identified, was initially described as being expressed in mature dendritic cells (DCs). Recent studies have shown that LAMP3 is also highly expressed in human lung tissue, particularly in type 2 alveolar epithelial cells (AEC2), and may be involved in the processing and/or transport of pulmonary surfactant[36],[37]. A proteomics study comparing patients with COVID-19 and idiopathic pulmonary fibrosis (IPF) revealed elevated levels of the alveolar type 2 cell marker LAMP3 in the serum of patients with severe atelectasis due to COVID-19; similarly, LAMP3 levels were elevated in IPF patients and showed a similar correlation with lung function[38], highlighting the potential value of LAMP3 as a biomarker of pulmonary remodeling.

WFDC2, also known as human epididymis protein 4 (HE4), is believed to be involved in the pathobiology of fibrotic diseases. Notably, WFDC2 has been described as being specifically expressed in activated fibroblasts and mediates renal fibrosis as a pan-protease inhibitor. Although the specific function of WFDC2 remains unclear, it inhibits the activity of multiple matrix metalloproteinases and specifically suppresses their contribution to the degradation of collagen I, suggesting that WFDC2 plays a functional role in the pathogenesis of fibrosis[39],[40]. In recent years, studies have evaluated the performance of WFDC2 as a candidate biomarker for the diagnosis of connective tissue disease-associated ILD. For instances, patients with idiopathic inflammatory myopathy who exhibit elevated WFDC2 levels have a higher prevalence of ILD[41]; serum WFDC2 levels in RA-ILD patients are elevated and closely correlated with ILD severity[42]; serum WFDC2 levels in patients with systemic sclerosis (SSc)-associated ILD are higher than in SSc patients without ILD, and pulmonary function parameters are negatively correlated with WFDC2 levels, supporting the potential clinical value of WFDC2 as a novel diagnostic aid for CTD-ILD patients[43]. In addition, previous studies have shown that markers such as LAG3, SPINT1, TNFRSF8, CD80, and CXCL13 are associated with CTD-ILD[44]–[48], although research on the predictive value of these proteins in CTD-ILD remains limited at present.

There is currently a lack of research utilizing plasma proteomics data to predict the onset of ILD, particularly in RA patients, and the lack of model interpretability limits their clinical utility. Machine learning models offer significant advantages in identifying complex patterns within clinical data, thereby enhancing predictive accuracy and practical application value. This study compared the performance of various models using ROC and DCA curves, ultimately selecting the CatBoost model for further analysis. Based on SHAP analysis, the CatBoost model demonstrated both strong predictive performance and enhanced interpretability in identifying the risk of ILD in RA patients[49]. However, this study has several limitations. First, the UK Biobank cohort and GWAS data were derived from European populations, and the generalizability of the findings to other populations requires further validation. Second, the definitions of RA and ILD were based on ICD-10 records and self-reports, which may introduce bias. Third, this study evaluated only plasma protein levels and did not cover the entire human proteome. Fourth, internal validation was conducted solely on the study dataset, and external validation using additional datasets was not performed.

## Data Availability

All data produced in the present study are available upon reasonable request to the authors

## Supplementary material

(Supplementary material comprises of data (including text), tables, and figures, and should be referenced in the following format: Supplementary Data S1, Supplementary Table S1, Supplementary Figure S1.)

