## Supplementary materials for "Development of a Novel Risk Prediction Model for Rheumatoid Arthritis–Associated Interstitial Lung Disease (RA-ILD): A Longitudinal Study"

Background: Interstitial lung disease (ILD) is one of the most common and potentially most devastating extra-articular complication of rheumatoid arthritis (RA) and is associated with substantial morbidity and mortality. However, reliable tools for the early identification of ILD in patients with RA remain limited. This study aimed to identify plasma protein biomarkers of RA-ILD and develop an interpretable machine learning model for risk prediction using data from the UK Biobank.

Methods: We first evaluated the association between baseline RA and the risk of incident ILD in the UK Biobank using Cox proportional hazards models. Mendelian randomization analysis was then performed to investigate the potential causal relationship between RA and ILD. Finally, we analyzed 2,920 plasma proteins measured using the Olink platform in 781 eligible RA patients. Proteins associated with ILD risk were identified using Cox proportional hazards models and subsequently used to construct eight machine learning models. Model performance was assessed using the receiver operating characteristic curve (ROC) and decision curve analysis. The best-performing model was further interpreted using Shapley additive explanations (SHAP) to evaluate feature importance.

Results: Compared with participants without RA, Patients with baseline RA had a significantly higher risk of developing ILD (Hazard ratio: 4.425, 95% CI: 3.549,5.518). The MR supported a potential causal association between RA and ILD (Odds ratio: 1.227, 95% CI: 1.121,1.343). Among the eight machine learning models, the CatBoost model showed the best performance, achieving an area under the curve (AUC) of 0.884 (95% CI: 0.773,0.996). The SHAP analysis identified LAG3, NPC2, and LAMP3 are the three most important plasma protein predictors of ILD development in patients with RA.

Conclusion: Plasma proteomics combined with machine learning may provide a promising approach for identifying biomarkers and predicting ILD risk in patients with RA. LAG3, NPC2, and LAMP3 may serve as candidate biomarkers for RA-ILD and warrant further validation.

Keywords: Rheumatoid arthritis, Interstitial lung disease, Mendelian randomization, Machine learning, Plasma proteins.

**Supplementary materials**

**Supplementary Table 1** Diet component definitions

**Supplementary Table 2** Baseline characteristics of matched participants

**Supplementary Table 3** SNPs used as instruments and their association with the exposure and outcome

**Supplementary Table 4** Measures of heterogeneity with Cochran’s Q test

**Supplementary Table 5** Examination of horizontal pleiotropy effects with MR-Egger regression tests

**Supplementary Table 6** Performance comparison of eight machine learning model in the training sets

**Supplementary Table 7** Performance comparison of eight machine learning model in the test sets

**Supplementary Figure 1** Flow chart of participants selection

**Supplementary Figure 2** Forest plot of multivariate Cox regression analysis

**Supplementary Figure 3** Scatter plot for the potential impacts of SNPs on both RA and ILD

**Supplementary Figure 4** Funnel plot for the causal relationship between RA and ILD

**Supplementary Figure 5** Forest plots of the leave-one-out analysis for the causal relationship between RA and ILD

**Supplementary Table 1** Diet component definitions

| Diet component | Field IDs | Amount per serving |
| --- | --- | --- |
| Fruit | 1309 (pieces fresh fruit/day)  1319 (pieces dried fruit/day) | 1309–1 piece  1319–5 pieces |
| Vegetable | 1289 (tablespoons cooked vegetables/day)  1299 (salad/raw vegetables/day) | 3 heaped tablespoons |
| (Shell)fish | 1329 (oily fish/week)  1339 (non-oily fish/week) | Once/week |
| Unprocessed meats | 1369 (beef/week or day)  1379 (lamb or mutton/week or day)  1389 (pork/week or day) | 1369-138 - once/week |
| Processed meats | 1349 (processed meat/week or daily) | 1349–1 piece/day |

Healthy diet was defined as satisfying ≥ 2 of the 4 criteria: (a) fruits/vegetables(including fresh fruit, dried fruit, cooked vegetables, and raw vegetables)> 4.5 servings/week; (b) fish(including oily fish and non-oily fish) >2 times/week; (c) processed meat ≤ 2 times/week; (d) red meat(including beef, lamb, and pork) < 5 times/week

**Supplementary Table 2** Baseline characteristics of matched participants

|  | Total | Non-RA | RA | *P*-value |
| --- | --- | --- | --- | --- |
| No.of participants | 24981 | 19980 | 5001 |  |
| Age,y | 58.71 (7.41) | 58.73 (7.46) | 58.66 (7.20) | 0.536 |
| Sex |  |  |  | 0.64 |
| Female | 17877 (71.6) | 14312 (71.6) | 3565 (71.3) |  |
| Male | 7104 (28.4) | 5668 (28.4) | 1436 (28.7) |  |
| Ethnicity |  |  |  | 0.988 |
| White | 22953 (91.9) | 18349 (91.8) | 4604 (92.1) |  |
| Mixed | 947 ( 3.8) | 765 ( 3.8) | 182 ( 3.6) |  |
| Asian | 671 ( 2.7) | 537 ( 2.7) | 134 ( 2.7) |  |
| Black | 152 ( 0.6) | 121 ( 0.6) | 31 ( 0.6) |  |
| Chinese | 51 ( 0.2) | 42 ( 0.2) | 9 ( 0.2) |  |
| Others | 207 ( 0.8) | 166 ( 0.8) | 41 ( 0.8) |  |
| Income |  |  |  | 0.712 |
| Less than £18,000 | 8834 (35.4) | 7053 (35.3) | 1781 (35.6) |  |
| £18,000 to £30,999 | 7246 (29.0) | 5816 (29.1) | 1430 (28.6) |  |
| £31,000 to £51,999 | 5264 (21.1) | 4228 (21.2) | 1036 (20.7) |  |
| £52,000to £100,000 | 3004 (12.0) | 2378 (11.9) | 626 (12.5) |  |
| Greater than £100,000 | 633 ( 2.5) | 505 ( 2.5) | 128 ( 2.6) |  |
| Education |  |  |  | 0.794 |
| College or University degree | 6722 (26.9) | 5366 (26.9) | 1356 (27.1) |  |
| Not-College or University degree | 12495 (50.0) | 10015 (50.1) | 2480 (49.6) |  |
| None of the above | 5764 (23.1) | 4599 (23.0) | 1165 (23.3) |  |
| TDI | -0.95 (3.22) | -0.96 (3.21) | -0.94 (3.25) | 0.693 |
| BMI,kg/m² | 28.23 (5.30) | 28.22 (5.26) | 28.30 (5.46) | 0.312 |
| Smoke |  |  |  | 0.945 |
| Never | 12050 (48.2) | 9628 (48.2) | 2422 (48.4) |  |
| Previous | 10116 (40.5) | 8101 (40.5) | 2015 (40.3) |  |
| Current | 2815 (11.3) | 2251 (11.3) | 564 (11.3) |  |
| Alcohol |  |  |  | 0.361 |
| Never | 1715 ( 6.9) | 1370 ( 6.9) | 345 ( 6.9) |  |
| Previous | 1730 ( 6.9) | 1361 ( 6.8) | 369 ( 7.4) |  |
| Current | 21536 (86.2) | 17249 (86.3) | 4287 (85.7) |  |
| Physical activities |  |  |  | 0.653 |
| No | 6250 (25.0) | 4986 (25.0) | 1264 (25.3) |  |
| Yes | 18731 (75.0) | 14994 (75.0) | 3737 (74.7) |  |
| Healthy diet |  |  |  | 0.461 |
| No | 4330 (17.3) | 3445 (17.2) | 885 (17.7) |  |
| Yes | 20651 (82.7) | 16535 (82.8) | 4116 (82.3) |  |
| HDL,mmol/L | 1.47 (0.38) | 1.47 (0.38) | 1.45 (0.38) | <0.001 |
| TG,mmol/L | 1.77 (1.01) | 1.77 (1.02) | 1.75 (1.00) | 0.212 |
| Glucose,mmol/L | 5.16 (1.23) | 5.16 (1.23) | 5.18 (1.25) | 0.302 |
| HbA1c,mmol/L | 36.50 (6.74) | 36.69 (6.65) | 35.76 (7.03) | <0.001 |
| ILD |  |  |  | <0.001 |
| No | 24660 (98.7) | 19825 (99.2) | 4835 (96.7) |  |
| Yes | 321 ( 1.3) | 155 ( 0.8) | 166 ( 3.3) |  |
| Follow-up time,m | 166.35 (13.70) | 166.55 (12.16) | 165.53 (18.61) | <0.001 |

Abbreviations:RA, Rheumatoid arthritis; TDI, Townsend deprivation index; BMI, body mass index; HDL, high density lipoprotein; TG, triglycerides; HbA1c, glycosylated hemoglobin; ILD, Interstitial lung disease; y, year; m, month.

**Supplementary Table 3** SNPs used as instruments and their association with the exposure and outcome

| SNP ID | EA | OA | EAF | Exposure | | | | Outcome | | |
| --- | --- | --- | --- | --- | --- | --- | --- | --- | --- | --- |
|  |  |  |  | *β* | SE | *P* value | F statistic | *β* | SE | *P* value |
| rs10261758 | A | G | 0.7107 | -0.081 | 0.0143 | 1.47×10^-08^ | 32.08 | -0.1043 | 0.0277 | 1.69×10^-04^ |
| rs10797431 | T | G | 0.4036 | -0.074 | 0.0129 | 1.06×10^-08^ | 32.91 | -0.0034 | 0.0254 | 8.94×10^-01^ |
| rs11889341 | T | C | 0.2496 | 0.1232 | 0.0146 | 2.71×10^-17^ | 71.21 | 0.0452 | 0.0288 | 1.17×10^-01^ |
| rs142144003 | A | G | 0.1112 | 0.4134 | 0.0207 | 1.09×10^-88^ | 398.84 | 0.1106 | 0.0429 | 9.93×10^-03^ |
| rs1611318 | C | A | 0.7616 | 0.1259 | 0.0185 | 1.07×10^-11^ | 46.31 | -0.0174 | 0.0376 | 6.45×10^-01^ |
| rs1776616 | G | A | 0.665 | 0.0832 | 0.0139 | 2.41×10^-09^ | 35.83 | -0.0135 | 0.0277 | 6.24×10^-01^ |
| rs2156698 | A | G | 0.4561 | -0.0745 | 0.0131 | 1.44×10^-08^ | 32.34 | 0.0071 | 0.0256 | 7.82×10^-01^ |
| rs2618444 | C | A | 0.3874 | 0.1005 | 0.0146 | 6.71×10^-12^ | 47.38 | 0.0396 | 0.0287 | 1.68×10^-01^ |
| rs28362859 | C | A | 0.1 | 0.1465 | 0.0205 | 9.90×10^-13^ | 51.07 | 0.0526 | 0.0421 | 2.11×10^-01^ |
| rs34536443 | C | G | 0.0428 | -0.2367 | 0.0427 | 3.03×10^-08^ | 30.73 | -0.1582 | 0.0785 | 4.38×10^-02^ |
| rs35139284 | T | C | 0.3191 | 0.5387 | 0.0149 | 1.00×10^-200^ | 1307.13 | 0.077 | 0.0289 | 7.60×10^-03^ |
| rs35511257 | C | G | 0.0746 | 0.6889 | 0.0408 | 4.38×10^-64^ | 285.10 | 0.0578 | 0.0712 | 4.17×10^-01^ |
| rs362531 | A | G | 0.0615 | -0.2121 | 0.0252 | 3.47×10^-17^ | 70.84 | -0.0023 | 0.0541 | 9.66×10^-01^ |
| rs3757387 | C | T | 0.3444 | 0.1086 | 0.0147 | 1.47×10^-13^ | 54.58 | 0.0774 | 0.0279 | 5.54×10^-03^ |
| rs56376587 | C | A | 0.4293 | 0.0785 | 0.0133 | 3.77×10^-09^ | 34.84 | 0.0265 | 0.026 | 3.07×10^-01^ |
| rs5754100 | C | T | 0.2441 | 0.086 | 0.015 | 9.20×10^-09^ | 32.87 | 0.0345 | 0.0295 | 2.42×10^-01^ |
| rs5757628 | A | G | 0.4275 | 0.0966 | 0.0157 | 7.58×10^-10^ | 37.86 | -0.0767 | 0.0303 | 1.13×10^-02^ |
| rs62395272 | T | C | 0.111 | 0.3146 | 0.0206 | 6.60×10^-53^ | 233.23 | 0.1178 | 0.0405 | 3.61×10^-03^ |
| rs6679677 | A | C | 0.1129 | 0.3051 | 0.0254 | 2.54×10^-33^ | 144.28 | 0.07 | 0.0461 | 1.29×10^-01^ |
| rs71508903 | T | C | 0.2109 | 0.0872 | 0.0157 | 2.54×10^-08^ | 30.85 | 0.0561 | 0.031 | 7.01×10^-02^ |
| rs7731626 | A | G | 0.2789 | -0.1343 | 0.0157 | 9.77×10^-18^ | 73.17 | -0.0327 | 0.0297 | 2.71×10^-01^ |
| rs9277814 | G | A | 0.5102 | 0.1322 | 0.0145 | 6.12×10^-20^ | 83.12 | 0.021 | 0.0279 | 4.50×10^-01^ |
| rs9469053 | G | A | 0.0454 | -0.2907 | 0.0312 | 1.22×10^-20^ | 86.81 | -0.0487 | 0.0628 | 4.38×10^-01^ |
| rs9494894 | C | T | 0.0417 | 0.1726 | 0.031 | 2.48×10^-08^ | 31.00 | 0.1728 | 0.0635 | 6.54×10^-03^ |

Abbreviations: EA, effect allele; EAF, effect allele frequency; MR, Mendelian randomization; OA, other allele; SE, standard error; β, effect estimate.

**Supplementary Table 4** Measures of heterogeneity with Cochran’s Q test

| Exposure | Outcome | Method | Q | Q_df | Q_pval |
| --- | --- | --- | --- | --- | --- |
| RA | ILD | MR Egger | 41.385 | 22 | 0.007 |
|  |  | IVW | 42.956 | 23 | 0.007 |

Abbreviation: RA, Rheumatoid arthritis; ILD, Interstitial lung disease; IVW, inverse variance weighted; MR, Mendelian randomization; Q_df, degrees of freedom associated with the Cochran Q test of heterogeneity; Q_pval, P value of Q test for heterogeneity.

**Supplementary Table 5** Examination of horizontal pleiotropy effects with MR-Egger regression tests

| Exposure | Outcome | egger_intercept | SE | *P* value |
| --- | --- | --- | --- | --- |
| RA | ILD | 0.01307 | 0.01430 | 0.37069 |

Abbreviation: RA, Rheumatoid arthritis; ILD, Interstitial lung disease; SE, standard error.

**Supplementary Table 6** Performance comparison of eight machine learning model in the training sets

| Model | Threshold | Accuracy | Sensitivity | Specificity | Precision | F1 |
| --- | --- | --- | --- | --- | --- | --- |
| Logistic | 0.661 | 0.894 | 0.846 | 0.942 | 0.936 | 0.86 |
| SVM | 0.655 | 0.885 | 0.808 | 0.962 | 0.955 | 0.833 |
| GBM | 0.505 | 0.885 | 0.865 | 0.904 | 0.9 | 0.87 |
| NeuralNetwork | 0.491 | 0.904 | 0.904 | 0.904 | 0.904 | 0.904 |
| Xgboost | 0.598 | 0.894 | 0.788 | 1 | 1 | 0.825 |
| Adaboost | 0.500 | 0.904 | 0.962 | 0.846 | 0.862 | 0.957 |
| LightGBM | 0.496 | 0.875 | 0.846 | 0.904 | 0.898 | 0.855 |
| CatBoost | 0.624 | 0.913 | 0.904 | 0.923 | 0.922 | 0.906 |

**Supplementary Table 7** Performance comparison of eight machine learning model in the test sets

| Model | Threshold | Accuracy | Sensitivity | Specificity | Precision | F1 |
| --- | --- | --- | --- | --- | --- | --- |
| Logistic | 0.661 | 0.727 | 0.773 | 0.682 | 0.708 | 0.75 |
| SVM | 0.655 | 0.773 | 0.818 | 0.727 | 0.75 | 0.8 |
| GBM | 0.505 | 0.682 | 0.864 | 0.5 | 0.633 | 0.786 |
| NeuralNetwork | 0.491 | 0.773 | 0.864 | 0.682 | 0.731 | 0.833 |
| Xgboost | 0.598 | 0.773 | 0.773 | 0.773 | 0.773 | 0.773 |
| Adaboost | 0.500 | 0.727 | 0.864 | 0.591 | 0.679 | 0.812 |
| LightGBM | 0.496 | 0.636 | 0.864 | 0.409 | 0.594 | 0.75 |
| CatBoost | 0.624 | 0.659 | 0.864 | 0.455 | 0.613 | 0.769 |

**Supplementary Figure 1** Flow chart of participants selection

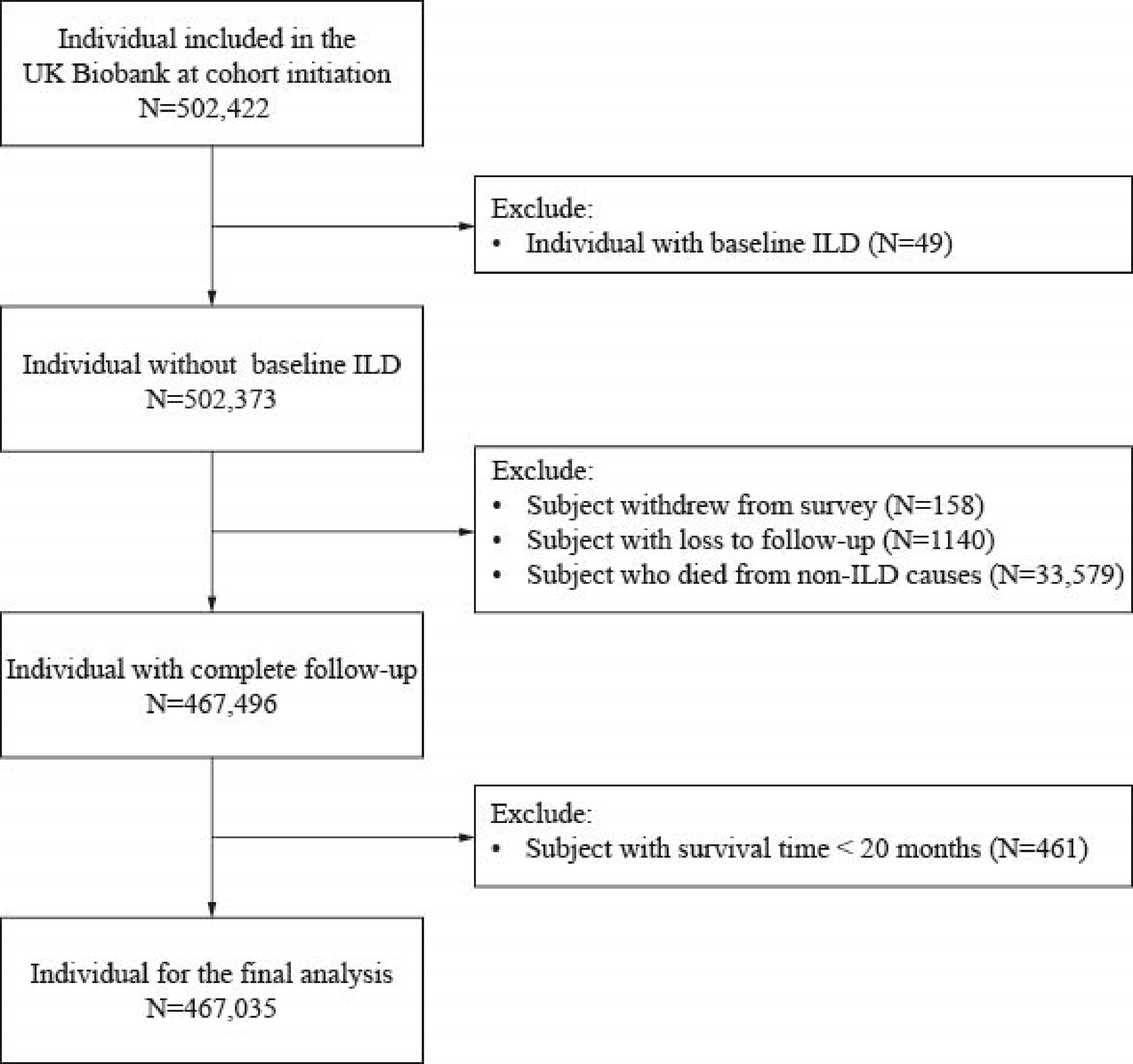

**Supplementary Figure 2** Forest plot of multivariate Cox regression analysis

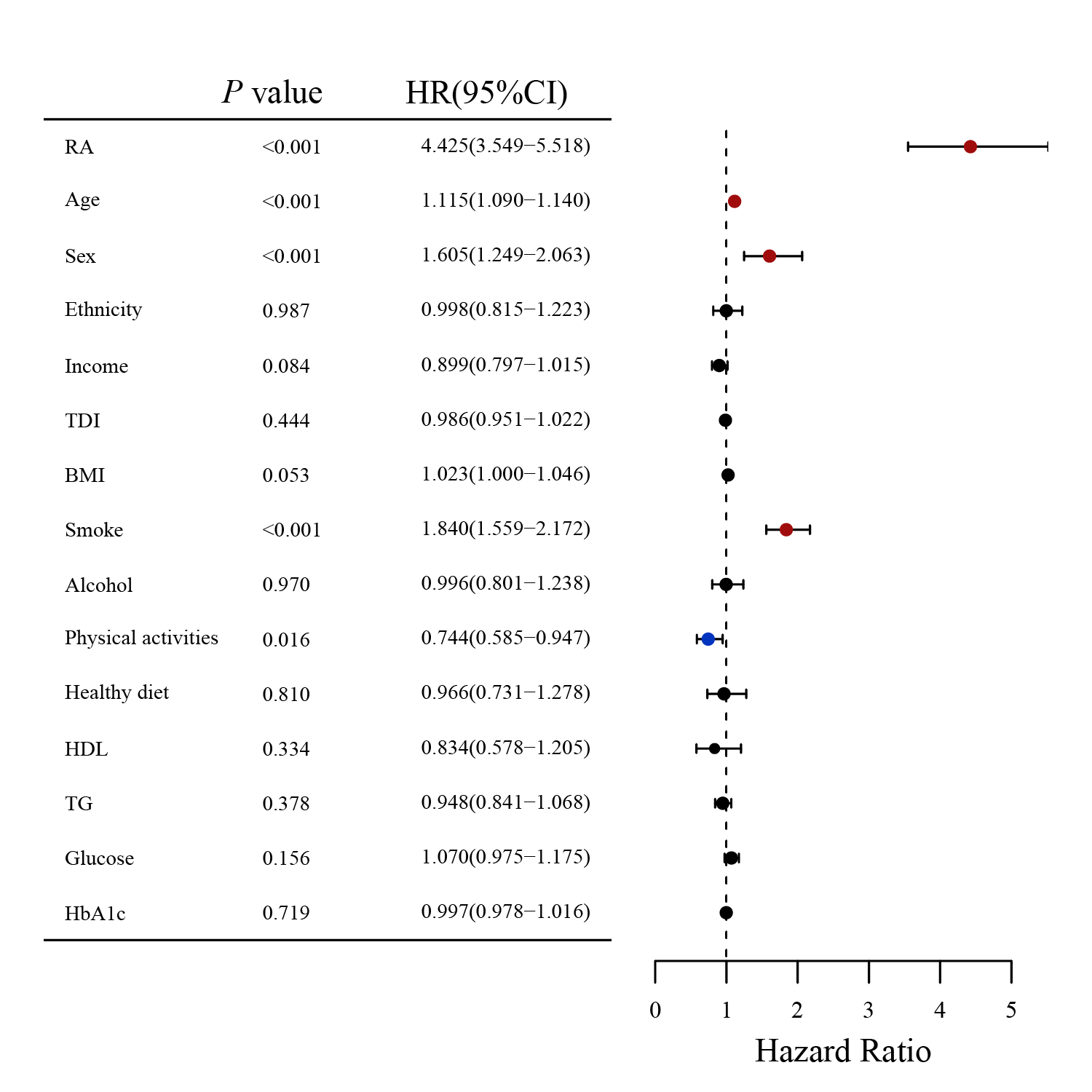

**Supplementary Figure 3** Scatter plot for the potential impacts of SNPs on both RA and ILD

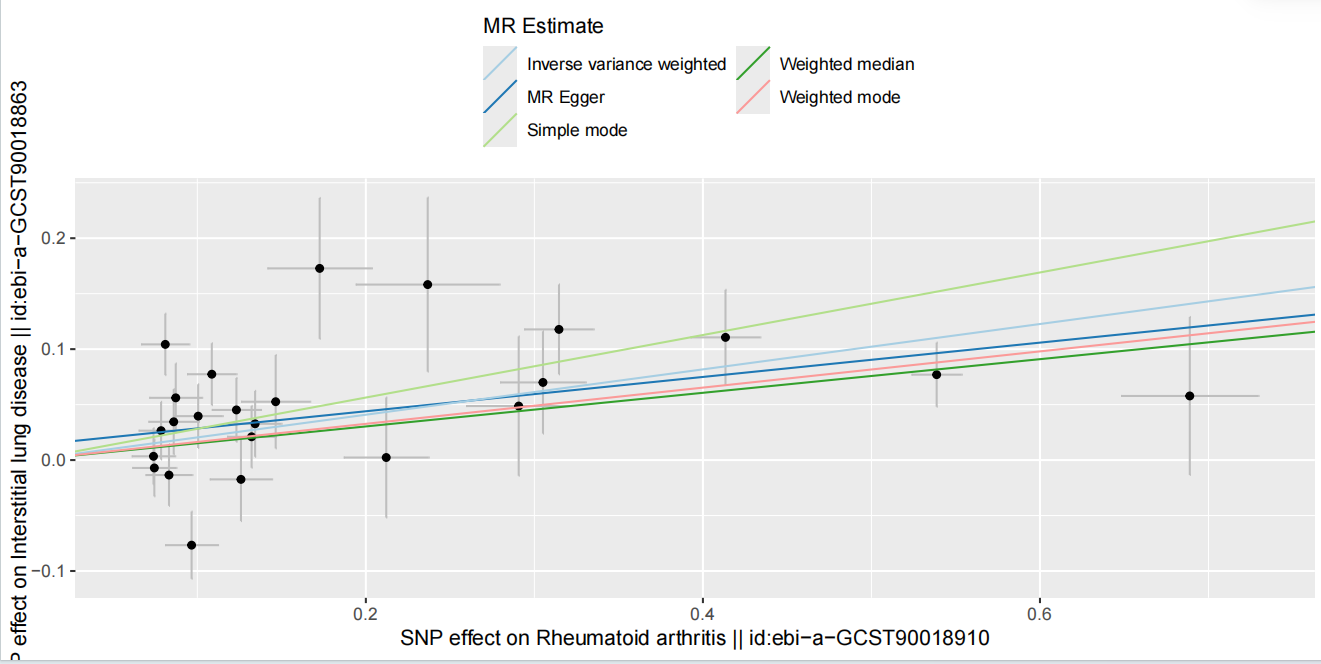

**Supplementary Figure 4** Funnel plot for the causal relationship between RA and ILD

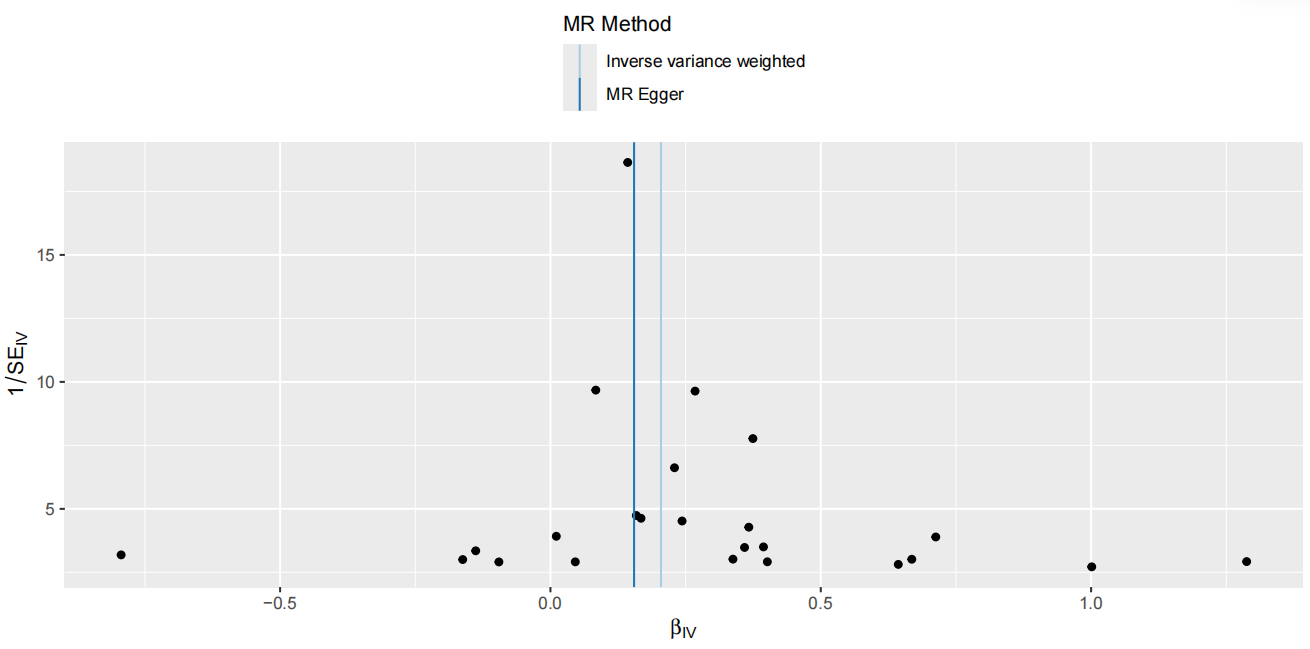

**Supplementary Figure 5** Forest plots of the leave-one-out analysis for the causal relationship between RA and ILD

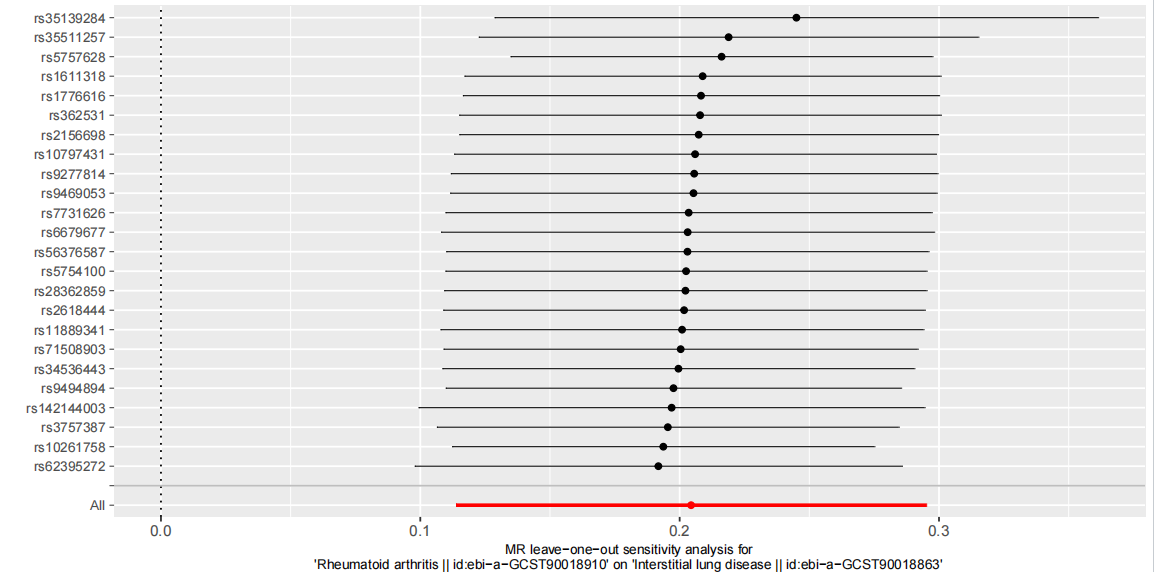
